# Impact of HPV vaccination on high-risk HPV prevalence in 25-29-year-old women attending cervical screening in England 2016-2025

**DOI:** 10.64898/2026.09.28.26364150

**Authors:** Lucinda Slater, Kavita Panwar, Marta Checchi, Laura Viviani, Alex Sargent, Viki Frew, Bethan Mackenzie, Simon Beddows, Kate Soldan

## Abstract

**Background:** In England, a national school-based human papillomavirus (HPV) vaccination programme was introduced in September 2008. Routine vaccination against HPV16/18 is offered to girls aged 12-13 and a ‘catch-up’ offer ran for girls aged 14-18 during the programme’s first two years.

**Methods:** Data from 19,407 women aged 25-29yrs attending cervical screening across England 2016-2025 were collected and residual samples from 4,837 high-risk (HR) HPV-screening-positive women were genotyped to monitor type-specific HR-HPV prevalence and evaluate vaccination impact.

**Results:** Type-specific HR-HPV results were available for 3,683 HR-HPV-screen-positive samples. Estimated age-adjusted prevalence of HPV16 and HPV18 was 91% (95% confidence interval [CI]: 87-94%) and 84% (95%CI: 65-93%) lower respectively among routine-eligible women than vaccination-ineligible women. Large declines were also observed for HPV31 (85% [95%CI: 72-92%]) and HPV45 (84% [95%CI: 73-90%]) but not for other HR-HPV types. Among 618 women with cytological abnormalities, HPV16/18 was present in samples from 53.9% (95%CI: 34.8-73.1%) of vaccination-ineligible, 18.1% (95%CI: 13.5-22.7%) of catch-up-eligible, and 6.5% (95%CI: 3.6-9.3%) of routine-eligible women.

**Conclusions:** The early years of the HPV vaccination programme in England conferred strong protection against HPV16, 18, 31 and 45, and dramatically changed the target remaining for screening strategies to detect and reduce cervical disease.

## Introduction

Human papillomavirus (HPV) is a large family of viruses, of which around 40 are sexually transmitted and infect the genital mucosa. Persistent infection with high-risk (HR) types is a necessary cause of cancers of the cervix, and a cause of some cancers of the anus, penis, vulva, vagina and oropharynx (1). Prior to widespread vaccination, two HR types, HPV16 and HPV18, were present in over 80% of all cervical cancers in the UK and in over 90% of those diagnosed in women under 30 years of age (2, 3).

In England, a national HPV vaccination programme began in September 2008, offering vaccination to adolescent girls via school-based delivery. The bivalent vaccine, which directly protects against HPV16 and HPV18, was offered to the first routine cohorts (born on or after 1^st^ September 1995) at age 12-13, with a catch-up offer for girls aged 14-18 (born on or after 1^st^ September 1990) during the first two years of the programme. The programme changed from September 2012 to offer a quadrivalent vaccine with additional direct protection against low-risk (LR) types HPV6 and HPV11 which cause 90% of genital warts (4). This was since changed to the nonavalent vaccine providing additional direct protection against HR-HPV types 31, 33, 45, 52, and 58 from 2022 (5). The programme was extended to include boys from 2019 (6).

Women who had been eligible for catch-up vaccination when aged 14-18 started to become eligible for the NHS Cervical Screening Programme (NHSCSP) at age 24.5 years in 2015, while those who had been eligible for routine vaccination at age 12-13 years started to become eligible in 2020 (7). In December 2019, the NHSCSP introduced HR-HPV testing as the primary screening test (8). If the HR-HPV screening test is positive (detects one or more of 13 HR HPV types: 16, 18, 31, 33, 35, 39, 45, 51, 52, 56, 58, 59, 68 (1) and dependent on testing platform, one possible HR type 66 (9)), samples proceed for a cytology screen for abnormal cells. These screening test results determine recall intervals and whether colposcopy and histology are indicated.

Surveillance of HPV infections amongst women aged 16-24 years attending chlamydia screening in England has shown reductions in the prevalence of HPV16/18 from 19.1% (95%CI: 16.6–21.8%) in 2008 to less than 1% (0.9%, 95%CI: 0.1-3.07%) in 2020 among 16–18-year-olds who would have been offered routine vaccination at age 12-13 years. Substantial, though smaller, declines were also seen for women offered catch-up vaccination at age 14-18 years (10, 11).

From 2007-2009 we conducted a baseline, pre-vaccination, survey of type-specific HR-HPV amongst women attending cervical screening (12). Here we report the first findings on the impact of routine vaccination on the cervical screening population in England, with data from a similar post-vaccination collection of samples from women aged 25 to 29 attending cervical screening from 2016-2025. The primary objectives of this surveillance were to (i) monitor trends in and estimate post-vaccination type-specific HPV prevalence among 25-29-year-old NHSCSP attendees by eligibility for HPV vaccination and (ii) to describe changes in the type-distribution of HR-HPV among women with abnormal cells detected during cytology, by eligibility for HPV vaccination.

## Methods

### Data and specimen collection

Data from a sample of 25-29-year-old women attending cervical screening between 2016-2025 were collected from a group of laboratories spread across England who participated in the pilot programme of HR-HPV primary screening (Southmead Hospital serving the South West, Norfolk and Norwich University Hospital serving the East of England, Northwick Park Hospital until 2020 and subsequently Health Services Laboratories serving London, and Manchester University NHS Foundation Trust serving Liverpool and Manchester). Each record was assigned a unique study identifier. Data including date of birth, date of attendance, NHS number and NHSCSP HR-HPV screening result, plus cytology data where available, were submitted for attendees without selecting for HR-HPV positivity to provide a random sample of those tested by the laboratory. Date of birth was used to assign vaccination cohort, and attendees were classified as either ineligible for vaccination (born 1^st^ September 1989-31^st^ August 1990), eligible for catch-up vaccination at age 14-18 (born 1^st^ September 1990-31^st^ August 1995) or eligible for routine vaccination at age 12-13 (born 1^st^ September 1995 to 31^st^ August 1999) (Table 1). Estimates of first-dose bivalent HPV vaccination coverage by cohort were taken from published data (Table 1) (13, 14).

**Table 1:** HPV vaccination cohorts, HPV vaccination eligibility and estimated coverage of those included in collection.

| HPV vaccination cohorts included in collection |  |  |  |  | Academic year (1<sup>st</sup> September- 31<sup>st</sup> August) |  |  |  |  |  |  |  |  |  |  |  |  |  |  |  |  |
| --- | --- | --- | --- | --- | --- | --- | --- | --- | --- | --- | --- | --- | --- | --- | --- | --- | --- | --- | --- | --- | --- |
| Vaccination eligibility group | Date of birth | Cohort # | No. women included in dataset (N= 19,407) | First-dose vaccination coverage (%) <sup>(13)</sup> <sup>(14)</sup> | 2008-2009 | 2009-2010 | 2010-2011 | 2011-2012 | 2012-2013 | 2013-2014 | 2014-2015 | 2015-2016 | 2016-2017 | 2017-2018 | 2018-2019 | 2019-2020 | 2020-2021 | 2021-2022 | 2022-2023 | 2023-2024 | 2024-2025 |
|  |  |  |  |  | Age at start of academic year |  |  |  |  |  |  | Sampling period |  |  |  |  |  |  |  |  |  |
| Ineligible | 01/09/1989-31/08/1990 | - | 1,189 | 0 | 18 | 19 | 20 | 21 | 22 | 23 | 24 | 25 | 26 | 27 | 28 | 29 | 30 | 31 | 32 | 33 | 34 |
| Catch-up | 01/09/1990-31/08/1991 | 2 | 2,537 | 66.1 | 17 | 18 | 19 | 20 | 21 | 22 | 23 | 24 | 25 | 26 | 27 | 28 | 29 | 30 | 31 | 32 | 33 |
|  | 01/09/1991-31/08/1992 | 3 | 1,472 | 55.6 | 16 | 17 | 18 | 19 | 20 | 21 | 22 | 23 | 24 | 25 | 26 | 27 | 28 | 29 | 30 | 31 | 32 |
|  | 01/09/1992-31/08/1993 | 4 | 1,805 | 59.8 | 15 | 16 | 17 | 18 | 19 | 20 | 21 | 22 | 23 | 24 | 25 | 26 | 27 | 28 | 29 | 30 | 31 |
|  | 01/09/1993-31/08/1994 | 5 | 2,578 | 78.4 | 14 | 15 | 16 | 17 | 18 | 19 | 20 | 21 | 22 | 23 | 24 | 25 | 26 | 27 | 28 | 29 | 30 |
|  | 01/09/1994-31/08/1995 | 6 | 2,593 | 81.9 | 13 | 14 | 15 | 16 | 17 | 18 | 19 | 20 | 21 | 22 | 23 | 24 | 25 | 26 | 27 | 28 | 29 |
| Routine | 01/09/1995-31/08/1996 | 1 | 2,654 | 89.4 | 12 | 13 | 14 | 15 | 16 | 17 | 18 | 19 | 20 | 21 | 22 | 23 | 24 | 25 | 26 | 27 | 28 |
|  | 01/09/1996-31/08/1997 | 7 | 2,081 | 85.9 | 11 | 12 | 13 | 14 | 15 | 16 | 17 | 18 | 19 | 20 | 21 | 22 | 23 | 24 | 25 | 26 | 27 |
|  | 01/09/1997-31/08/1998 | 8 | 1,416 | 88.9 | 10 | 11 | 12 | 13 | 14 | 15 | 16 | 17 | 18 | 19 | 20 | 21 | 22 | 23 | 24 | 25 | 26 |
|  | 01/09/1998-31/08/1999 | 9 | 1,082 | 90.6 | 9 | 10 | 11 | 12 | 13 | 14 | 15 | 16 | 17 | 18 | 19 | 20 | 21 | 22 | 23 | 24 | 25 |

Offered first dose bivalent HPV vaccination
NHSCSP attendance sampled (ages 25-29 eligible only)

For the sampled attendees with a HR-HPV-positive screening result (positive for any HR types 16, 18, 31, 33, 35, 39, 45, 51, 52, 56, 58, 59, 68, and/or possible HR type 66), residual liquid-based cytology (LBC) samples were sent to UKHSA with all patient identifiers removed but with the unique study identifier for the patient record attached. Corresponding cytology outcomes were requested and where available were added to the dataset. The residual LBC samples were genotyped using an in-house ISO 15189:2022 accredited multiplex PCR and Luminex-based genotyping assay for multiple HPV types including the 13 HR types (1) and 5 possible HR types (HPV types 26, 53, 66, 70 and 73). Personal identifiers (date of birth and NHS number) were deleted once vaccination category was determined and before test results were added to the anonymised data file. Data from samples which did not produce a HR-HPV result for the 13 confirmed HR types (inadequate samples, those with only HPV66, and those with no type-specific result from the UKHSA assay) were excluded from the final dataset. Samples were classified as positive or negative for the following groups based on the UKHSA test result: vaccine-types HPV16/18; types with established cross-protection from the bivalent vaccine HPV31/33/45 (15, 16); non-16/18 HR nonavalent vaccine types HPV31/33/45/52/58, and non-vaccine HR types HPV35/39/51/56/59/68. The combined prevalence of the 5 possible HR types (HPV26/53/66/70/73), which are not affected by vaccination, was used as an indicator of sexual risk among each HR-HPV positive type grouping where individuals had coinfections with at least one HR type and at least one of these five additional HPV types (17).

### Statistical analysis

Pearson’s chi^2^ (Χ^2^) was used to test differences between estimates of proportions and prevalence for categorical variables. Log binomial regression was used to assess differences in type-specific HR-HPV prevalence estimates between the catch-up-eligible versus the ineligible group, and secondly the routine-eligible group versus the ineligible group, producing crude prevalence ratios (PRs) and 95% confidence intervals (95%CI). A multivariable log binomial regression model was fitted to adjust for potential confounders, including age at attendance, submitting laboratory and sexual risk indicator, producing adjusted prevalence ratios (aPR).

To further quantify the association between first-dose HPV vaccination coverage and changes in HR-HPV prevalence, we included a continuous variable in the regression model with the first-dose HPV vaccination coverage as a proportion (Table 1). Vaccine effectiveness was calculated as 1 – aPR (18). This vaccine effectiveness estimate used national first-dose coverage as a likelihood of vaccination however most vaccinated individuals would have received two or three vaccine doses according to the early vaccination schedule for adolescent girls.

Type-specific HR-HPV prevalence estimates and 95%CI among ineligible, catch-up-eligible and routine-eligible women were calculated for each type grouping. These were age-adjusted using predicted margins from the log binomial regression model to account for different age distributions among the ineligible, catch-up-eligible and routine-eligible groups. Age-adjusted proportion of type-specific HR-HPV was also estimated among those with a finding of abnormal cells during cervical cytology where data were available.

### Sensitivity Analyses

An initial sensitivity analysis was performed to determine the effect of excluding data that tested HR-HPV screen positive, but that did not produce a type-specific HR-HPV result (N=1,154), and those testing positive for only HPV66 (N=160). The adjusted prevalence (adjusted for age at attendance, submitting laboratory and sexual risk indicator) among HR-HPV positive samples which did have a type-specific result was applied to the number of screening-positive samples excluded without a type-specific result to impute the estimated number of samples expected to test positive for each HPV type. For each HPV type and type grouping, this additional number of “positives” was added to the known HPV positive types and used as the numerator. HPV screen negatives plus those with only HPV66 present were considered negative for HR-HPV, with the total used as the denominator to produce updated prevalence estimates. The absolute difference in percentage-points was calculated for each prevalence estimate to descriptively compare outputs from each method.

A second sensitivity analysis was performed excluding data (N=1,938) submitted by some laboratories for years when selection was evidently not random and HR-HPV screening positive samples made up a significantly higher or lower proportion of the total data received compared with those from other laboratory sites in the same year. Signals of over or under-sampling were investigated and where a reason was identified, including collection interruptions during the COVID-19 pandemic and misunderstanding of sampling instructions, these data were excluded. Age-adjusted HR-HPV prevalence was estimated across this dataset and compared with the primary analysis dataset to determine whether these data reporting issues impacted the results of the analysis. The absolute difference in percentage-points was calculated for each prevalence estimate to descriptively compare outputs from each method. All analyses were completed using STATA v18.

## Results

### Data collected

A total of 20,561 records from women aged 25-29 attending cervical screening were collected from 2016 to 2025, with 19,407 records included in the final dataset. Of 4,837 eligible HR-HPV screening-positive samples sent to UKHSA, 160 were positive for possible HR type HPV66 only, so not considered HR and were excluded from further analysis. Of 4,677 remaining, HR-HPV type data were available for 3,683 (79%), and only those with a type-specific HR-HPV result were included in the final dataset as HR-HPV positive samples. Those excluded in addition to the HPV66 positives comprised 8 with inadequate samples for testing, 569 in which no HPV DNA was detected, and a further 417 which had no HR-HPV typing result (only other possible HR/ low-risk HPV types). Of those with a positive HR-HPV screen and HR-HPV genotype data available, cytology data were provided for 2,369, of which 2,315 (63% of all included HR-HPV positives) were valid cytology results. Approximately 25% (n=618) of these had a finding of abnormal cells. Data for an additional 15,724 HR-HPV screening negative samples were received and included in the final dataset and assumed to be HR-HPV negative for analysis. The HR-HPV-screen-positive data excluded from analysis was comparable to the typed HR-HPV positives included in the final dataset by age at attendance (Χ ^2^= 5.72, *p*=0.221) and by submitting laboratory (Χ ^2^= 4.982, *p*=0.418), but had a higher proportion of later sampling years (Χ ^2^= 34.931, *p*<0.001), and younger vaccination cohorts (Χ ^2^= 25.224, *p*=0.003) (Table 2).

**Table 2.**
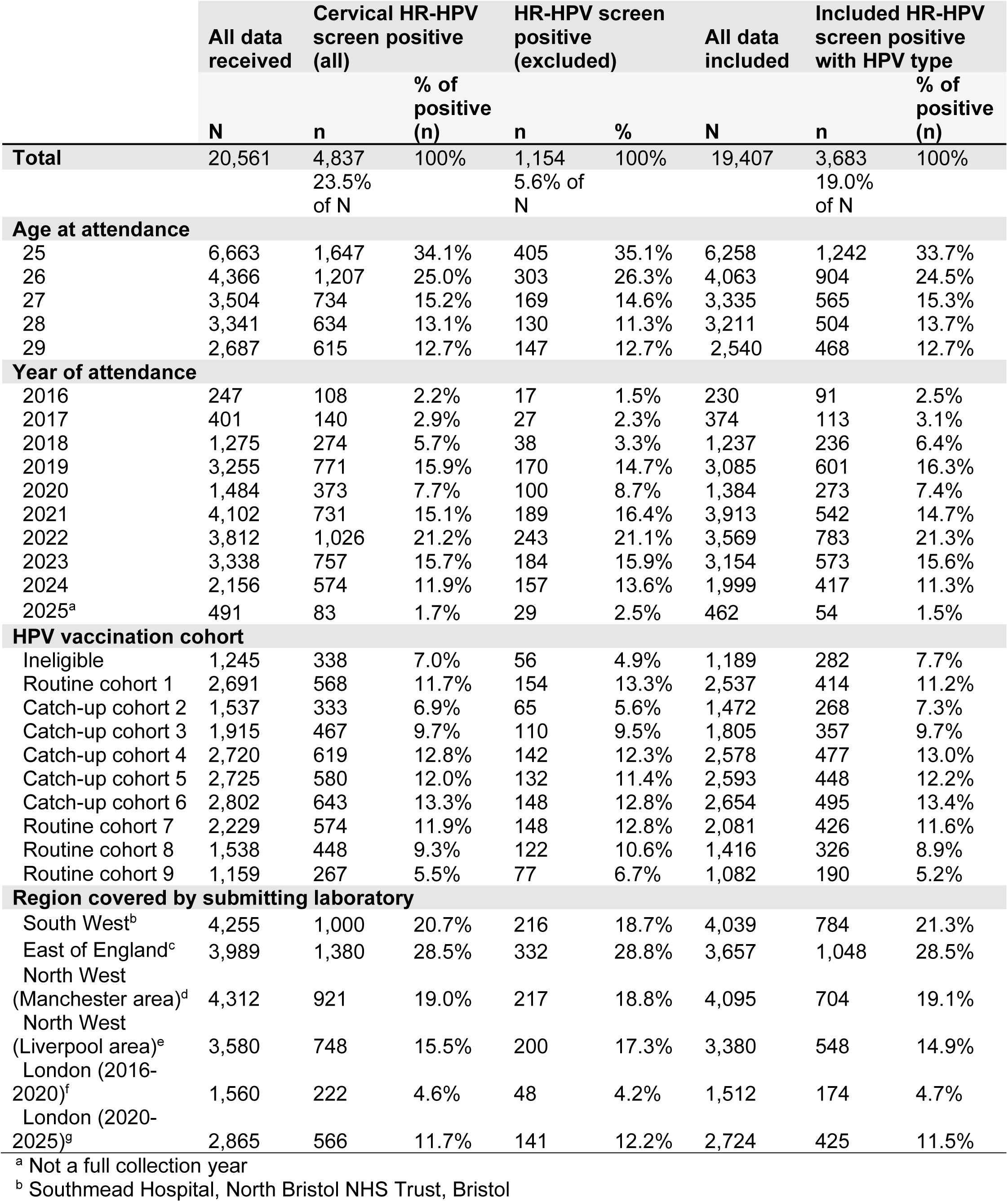
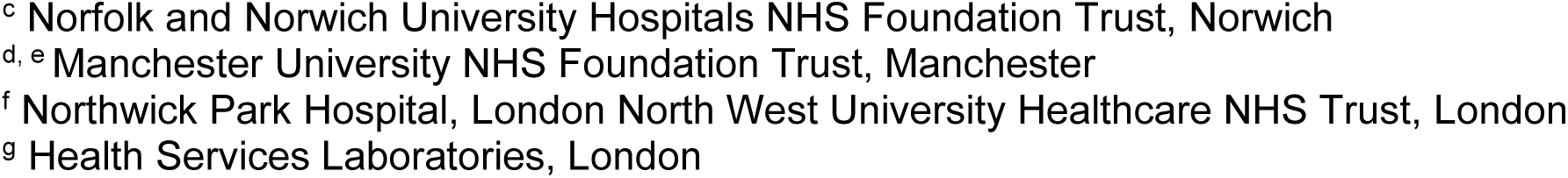
Summary of all data from 25-29-year-old women attending cervical screening 2016-2025 by age, year, vaccination cohort and submitting laboratory as a proportion of those received and those with a positive type-specific HR-HPV result include.

|  | All data received | Cervical HR-HPV screen positive (all) | % of positive (n) | HR-HPV screen positive (excluded) |  | All data included | Included HR-HPV screen positive with HPV type | % of positive (n) |
| --- | --- | --- | --- | --- | --- | --- | --- | --- |
|  | N | n |  | n | % | N | n |  |
| **Total** | 20,561 | 4,837 | 100% | 1,154 | 100% | 19,407 | 3,683 | 100% |
|  |  | 23.5% of N |  | 5.6% of N |  |  | 19.0% of N |  |
| **Age at attendance** |  |  |  |  |  |  |  |  |
| 25 | 6,663 | 1,647 | 34.1% | 405 | 35.1% | 6,258 | 1,242 | 33.7% |
| 26 | 4,366 | 1,207 | 25.0% | 303 | 26.3% | 4,063 | 904 | 24.5% |
| 27 | 3,504 | 734 | 15.2% | 169 | 14.6% | 3,335 | 565 | 15.3% |
| 28 | 3,341 | 634 | 13.1% | 130 | 11.3% | 3,211 | 504 | 13.7% |
| 29 | 2,687 | 615 | 12.7% | 147 | 12.7% | 2,540 | 468 | 12.7% |
| **Year of attendance** |  |  |  |  |  |  |  |  |
| 2016 | 247 | 108 | 2.2% | 17 | 1.5% | 230 | 91 | 2.5% |
| 2017 | 401 | 140 | 2.9% | 27 | 2.3% | 374 | 113 | 3.1% |
| 2018 | 1,275 | 274 | 5.7% | 38 | 3.3% | 1,237 | 236 | 6.4% |
| 2019 | 3,255 | 771 | 15.9% | 170 | 14.7% | 3,085 | 601 | 16.3% |
| 2020 | 1,484 | 373 | 7.7% | 100 | 8.7% | 1,384 | 273 | 7.4% |
| 2021 | 4,102 | 731 | 15.1% | 189 | 16.4% | 3,913 | 542 | 14.7% |
| 2022 | 3,812 | 1,026 | 21.2% | 243 | 21.1% | 3,569 | 783 | 21.3% |
| 2023 | 3,338 | 757 | 15.7% | 184 | 15.9% | 3,154 | 573 | 15.6% |
| 2024 | 2,156 | 574 | 11.9% | 157 | 13.6% | 1,999 | 417 | 11.3% |
| 2025<sup>a</sup> | 491 | 83 | 1.7% | 29 | 2.5% | 462 | 54 | 1.5% |
| **HPV vaccination cohort** |  |  |  |  |  |  |  |  |
| Ineligible | 1,245 | 338 | 7.0% | 56 | 4.9% | 1,189 | 282 | 7.7% |
| Routine cohort 1 | 2,691 | 568 | 11.7% | 154 | 13.3% | 2,537 | 414 | 11.2% |
| Catch-up cohort 2 | 1,537 | 333 | 6.9% | 65 | 5.6% | 1,472 | 268 | 7.3% |
| Catch-up cohort 3 | 1,915 | 467 | 9.7% | 110 | 9.5% | 1,805 | 357 | 9.7% |
| Catch-up cohort 4 | 2,720 | 619 | 12.8% | 142 | 12.3% | 2,578 | 477 | 13.0% |
| Catch-up cohort 5 | 2,725 | 580 | 12.0% | 132 | 11.4% | 2,593 | 448 | 12.2% |
| Catch-up cohort 6 | 2,802 | 643 | 13.3% | 148 | 12.8% | 2,654 | 495 | 13.4% |
| Routine cohort 7 | 2,229 | 574 | 11.9% | 148 | 12.8% | 2,081 | 426 | 11.6% |
| Routine cohort 8 | 1,538 | 448 | 9.3% | 122 | 10.6% | 1,416 | 326 | 8.9% |
| Routine cohort 9 | 1,159 | 267 | 5.5% | 77 | 6.7% | 1,082 | 190 | 5.2% |
| **Region covered by submitting laboratory** |  |  |  |  |  |  |  |  |
| South West<sup>b</sup> | 4,255 | 1,000 | 20.7% | 216 | 18.7% | 4,039 | 784 | 21.3% |
| East of England<sup>c</sup> | 3,989 | 1,380 | 28.5% | 332 | 28.8% | 3,657 | 1,048 | 28.5% |
| North West (Manchester area)<sup>d</sup> | 4,312 | 921 | 19.0% | 217 | 18.8% | 4,095 | 704 | 19.1% |
| North West (Liverpool area)<sup>e</sup> | 3,580 | 748 | 15.5% | 200 | 17.3% | 3,380 | 548 | 14.9% |
| London (2016-2020)<sup>f</sup> | 1,560 | 222 | 4.6% | 48 | 4.2% | 1,512 | 174 | 4.7% |
| London (2020-2025)<sup>g</sup> | 2,865 | 566 | 11.7% | 141 | 12.2% | 2,724 | 425 | 11.5% |

<sup>a</sup> Not a full collection year
<sup>b</sup> Southmead Hospital, North Bristol NHS Trust, Bristol
- <sup>c</sup> Norfolk and Norwich University Hospitals NHS Foundation Trust, Norwich - <sup>d, e</sup> Manchester University NHS Foundation Trust, Manchester - <sup>f</sup> Northwick Park Hospital, London North West University Healthcare NHS Trust, London - <sup>g</sup> Health Services Laboratories, London

In the final dataset, the overall mean age at attendance was 26.6 years (interquartile range [IQR]: 25-28). Older ages were overrepresented in the vaccination-ineligible (mean age 27.9 years, IQR: 27-29) and catch-up group (mean age: 27.0 years, IQR: 26-28) and underrepresented in the routine-eligible group (mean age: 25.7 years, IQR: 25-26), prior to age adjustment.

### Bivalent vaccine types (HPV16/18)

Estimated age-adjusted prevalence of HPV16 was 7.7% (95% confidence intervals [95%CI]: 5.7-11.4%) among the ineligible group, 2.1% (95%CI: 1.8-2.3%) among the catch-up-eligible group and 0.7% (95%CI: 0.5-0.8%) among the routine-eligible group (Table 3**Error! Reference source not found.**). Following further adjustment for age, submitting laboratory and sexual risk indicator, estimated HPV16 prevalence was 91% (95%CI: 87-94%) lower among those eligible for routine vaccination at age 12-13 and 72% (95%CI: 63-79%) lower among those eligible for catch-up vaccination compared with those ineligible for vaccination respectively (Figure 1; Table 4). Estimated age-adjusted prevalence of HPV18 was 1.7% (95%CI: 0.9-2.5%) among the ineligible group, 0.5% (95%CI: 0.4-0.7%) among the catch-up-eligible group and 0.2% (95%CI: 0.1-0.3%) among the routine-eligible group (Table 3). Following adjustment for confounders, estimated HPV18 prevalence was 84% (95%CI: 65-93%) lower among the routine-eligible group (aPR: 0.16, 95%CI: 0.07-0.35) and 63% (95%CI:36-78%) lower among the catch-up-eligible group (aPR: 0.37, 95%CI: 0.22-0.64) compared with the ineligible group (Figure 1; Table 4).

**Figure 1.**
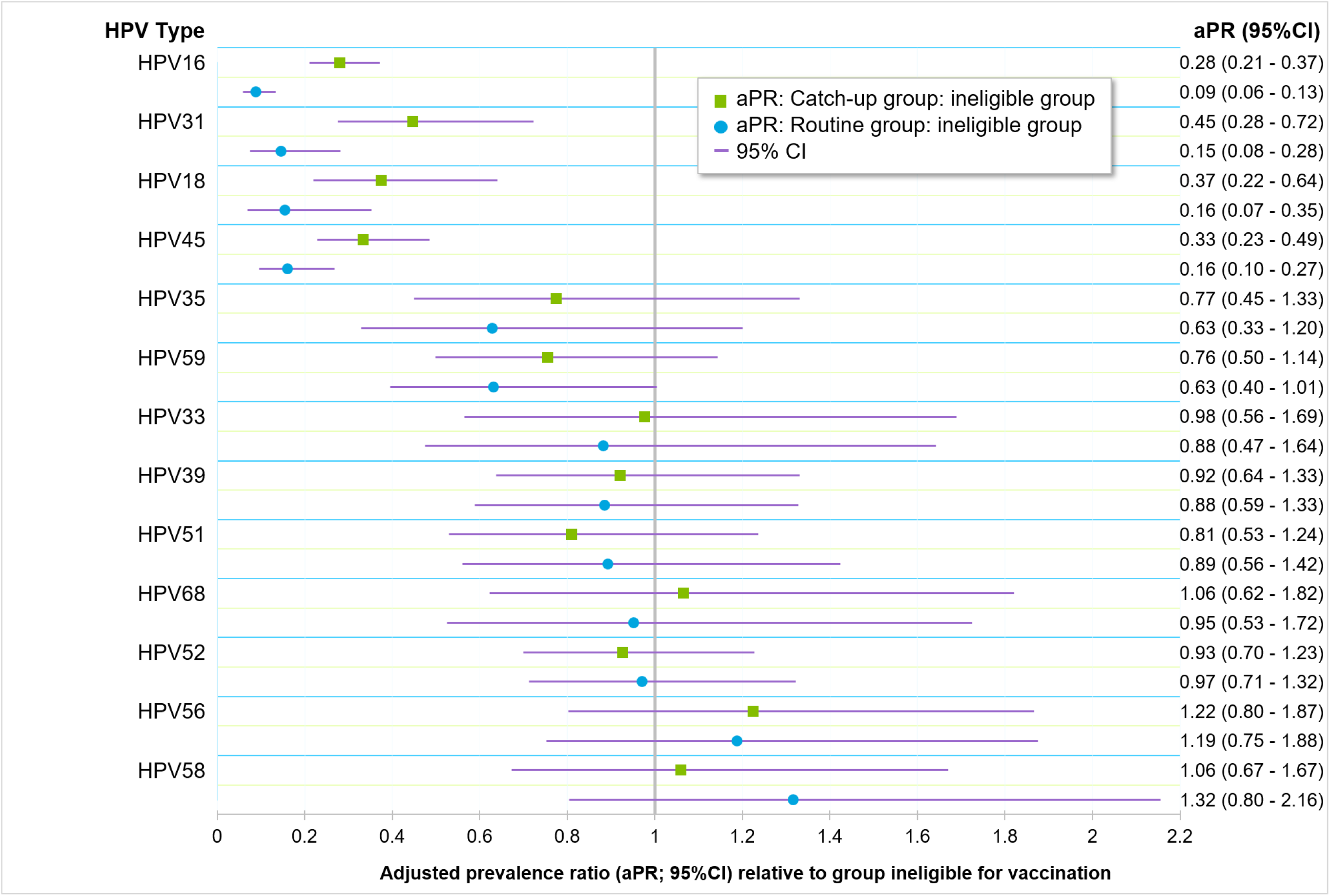
Adjusted prevalence ratio of type-specific HPV among women aged 25-29 attending cervical screening 2016-2025, by catch-up or routine eligibility for vaccination compared with vaccination-ineligible group, ordered by aPR of routine: ineligible group.

**Table 3.**
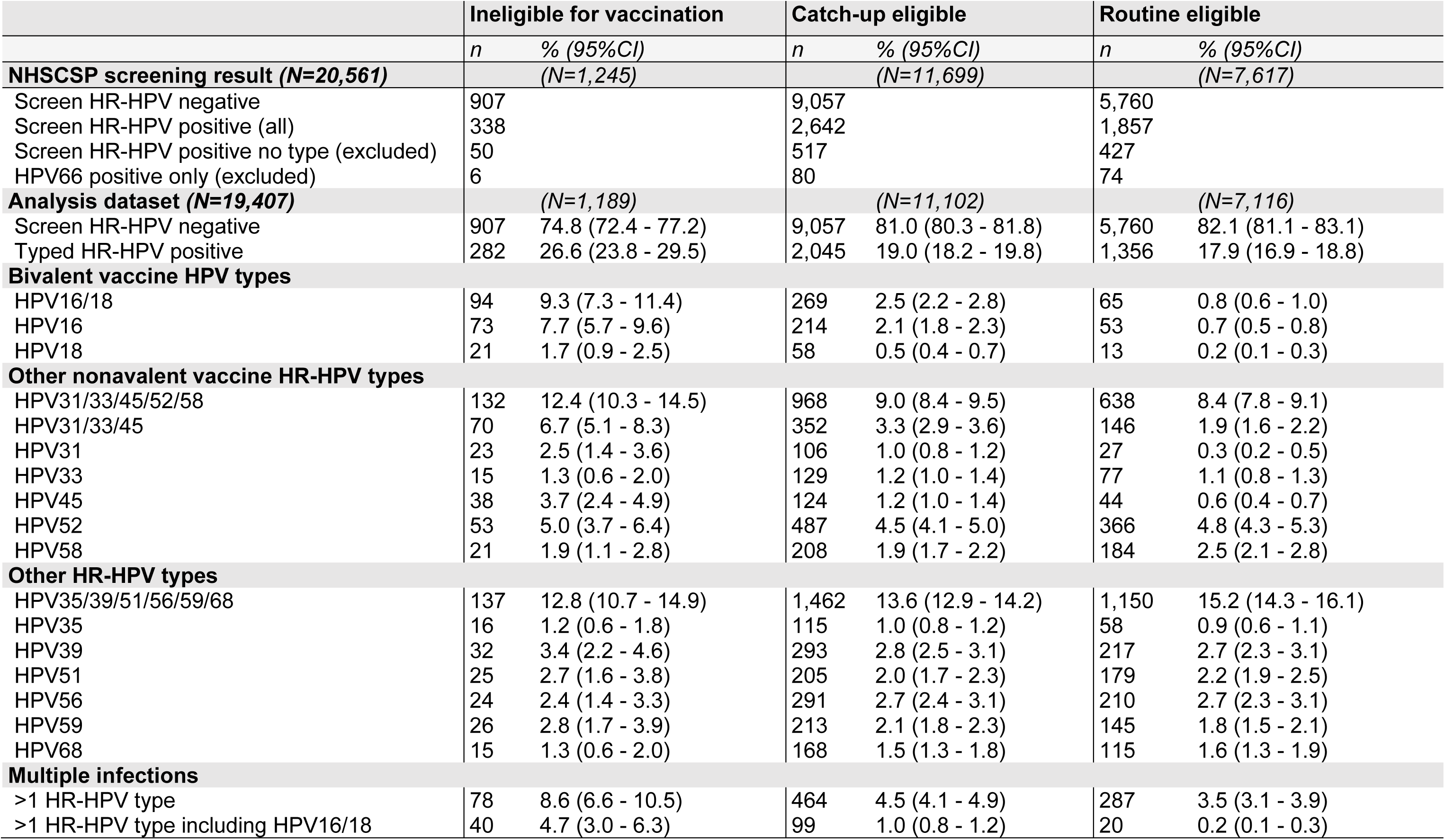
Estimated age-adjusted type-specific high-risk HPV prevalence among women attending for cervical screening, by vaccination cohort eligibility 2016-2025.

**Table 4.** Prevalence ratio of type-specific high-risk HPV among women attending for cervical screening 2016-2025, by vaccination eligibility and by estimated first-dose vaccination coverage.

|  | Catch-up eligible vs ineligible for vaccination |  | Routine eligible vs ineligible for vaccination |  | Prevalence ratio associated with first-dose vaccination coverage <sup>c</sup> |  |
| --- | --- | --- | --- | --- | --- | --- |
|  | *Unadjusted PR<sup>a</sup>*<br>(95%CI) | *Adjusted PR<sup>b</sup>*<br>(95%CI) | *Unadjusted PR*<br>(95%CI) | *Adjusted PR*<br>(95%CI) | *Unadjusted PR*<br>(95%CI) | *Adjusted PR*<br>(95%CI) |
| Any HR-HPV | **0.78 (0.70 - 0.87)** | **0.77 (0.68 - 0.87)** | **0.80 (0.72 - 0.90)** | **0.73 (0.64 - 0.84)** | 0.77 (0.57 - 1.04) | 0.97 (0.76 - 1.24) |
| **Bivalent vaccine HPV types** |  |  |  |  |  |  |
| HPV16 and/or 18 | **0.31 (0.24 - 0.38)** | **0.30 (0.23 - 0.38)** | **0.12 (0.08 - 0.16)** | **0.10 (0.07 - 0.14)** | **0.03 (0.01 - 0.06)** | **0.03 (0.01 - 0.07)** |
| HPV16 | **0.31 (0.24 - 0.41)** | **0.28 (0.21 - 0.37)** | **0.12 (0.09 - 0.17)** | **0.09 (0.06 - 0.13)** | **0.03 (0.01 - 0.08)** | **0.04 (0.02 - 0.10)** |
| HPV18 | **0.30 (0.18 - 0.49)** | **0.37 (0.22 - 0.64)** | **0.10 (0.05 - 0.21)** | **0.16 (0.07 - 0.35)** | **0.02 (<0.01- 0.13)** | **0.01 (<0.01- 0.08)** |
| **Other nonavalent vaccine HR-HPV types** |  |  |  |  |  |  |
| HPV31/33/45/52/58 | **0.79 (0.66 - 0.93)** | **0.78 (0.66 - 0.92)** | **0.81 (0.68 - 0.96)** | **0.75 (0.62 - 0.90)** | 0.78 (0.52 - 1.18) | 1.00 (0.69 - 1.46) |
| HPV31/33/45 | **0.54 (0.42 - 0.69)** | **0.54 (0.42 - 0.70)** | **0.35 (0.26 - 0.46)** | **0.32 (0.23 - 0.44)** | **0.09 (0.04 - 0.18)** | **0.12 (0.06 - 0.23)** |
| HPV31 | **0.49 (0.32 - 0.77)** | **0.45 (0.28 - 0.72)** | **0.20 (0.11 - 0.34)** | **0.15 (0.08 - 0.28)** | **0.01 (<0.01- 0.02)** | **0.01 (<0.01- 0.05)** |
| HPV33 | 0.92 (0.54 - 1.57) | 0.98 (0.56 - 1.69) | 0.86 (0.49 - 1.49) | 0.88 (0.47 - 1.64) | 0.83 (0.24 - 2.89) | 0.82 (0.28 - 2.45) |
| HPV45 | **0.35 (0.24 - 0.50)** | **0.33 (0.23 - 0.49)** | **0.19 (0.13 - 0.30)** | **0.16 (0.10 - 0.27)** | **0.04 (0.01 - 0.15)** | **0.07 (0.02 - 0.21)** |
| HPV52 | 0.98 (0.75 - 1.30) | 0.93 (0.70 - 1.23) | 1.15 (0.87 - 1.53) | 0.97 (0.71 - 1.32) | 1.47 (0.79 - 2.71) | **2.03 (1.18 - 3.50)** |
| HPV58 | 1.06 (0.68 - 1.65) | 1.06 (0.67 - 1.67) | 1.46 (0.94 - 2.29) | 1.32 (0.80 - 2.16) | **4.31 (1.63 - 11.42)** | **6.00 (2.53 - 14.20)** |
| **Other HR-HPV types** |  |  |  |  |  |  |
| HPV35 | 0.77 (0.46 - 1.29) | 0.77 (0.45 - 1.33) | 0.61 (0.35 - 1.05) | 0.63 (0.33 - 1.20) | **0.23 (0.06 - 0.89)** | **0.24 (0.08 - 0.76)** |
| HPV39 | 0.98 (0.68 - 1.41) | 0.92 (0.64 - 1.33) | 1.13 (0.79 - 1.63) | 0.88 (0.59 - 1.33) | 0.67 (0.31 - 1.47) | 1.31 (0.65 - 2.63) |
| HPV51 | 0.88 (0.58 - 1.32) | 0.81 (0.53 - 1.24) | 1.20 (0.79 - 1.81) | 0.89 (0.56 - 1.42) | 1.12 (0.44 - 2.87) | 2.65 (1.15 - 6.11) |
| HPV56 | 1.30 (0.86 - 1.96) | 1.22 (0.80 - 1.87) | 1.46 (0.96 - 2.22) | 1.19 (0.75 - 1.88) | 1.26 (0.56 - 2.82) | 1.94 (0.94 - 3.97) |
| HPV59 | 0.88 (0.59 - 1.31) | 0.76 (0.50 - 1.14) | 0.93 (0.62 - 1.41) | 0.63 (0.40 - 1.01) | 0.85 (0.32 - 2.22) | 1.74 (0.74 - 4.06) |
| HPV68 | 1.20 (0.71 - 2.03) | 1.06 (0.62 - 1.82) | 1.28 (0.75 - 2.19) | 0.95 (0.53 - 1.72) | 0.52 (0.18 - 1.54) | 1.06 (0.41 - 2.69) |
| **Multiple infections** |  |  |  |  |  |  |
| >1 high-risk HPV type | **0.64 (0.51 - 0.80)** | **0.55 (0.44 - 0.70)** | **0.61 (0.48 - 0.78)** | **0.42 (0.32 - 0.56)** | 0.36 (0.19 - 0.68) | 0.80 (0.46 - 1.40) |
| >1 high-risk HPV type including HPV16/18 | **0.27 (0.18 - 0.38)** | **0.22 (0.15 - 0.33)** | **0.08 (0.05 - 0.14)** | **0.05 (0.03 - 0.10)** | **0.02 (<0.01- 0.07)** | **0.03 (0.01 - 0.10)** |

<sup>a</sup> Prevalence ratio for catch-up or routine-eligible group versus baseline ineligible group.
<sup>b</sup> Adjusted prevalence ratio for catch-up or routine-eligible group versus baseline ineligible group, adjusted for age at attendance, submitting laboratory and sexual risk.
<sup>c</sup> Prevalence ratios for the association between first-dose HPV vaccination coverage by cohort and changes in estimated HPV prevalence.

Decreases in HPV16/18 prevalence estimates were strongly associated with first-dose HPV vaccination coverage (HPV16 aPR: 0.03 [95%CI: 0.01-0.06]; HPV18 aPR: 0.04 [95%CI: <0.01-0.08]), corresponding to an estimated vaccine effectiveness of 97% against HPV16/18 (aPR: 0.03 [95%CI: 0.01-0.07]; Table 4). While crude estimated HPV16/18 prevalence was highest among the younger years of age in the ineligible group (Χ ^2^= 36.2, *p*<0.001), this was no longer true among the catch-up (Χ ^2^= 6.8, *p*=0.147) or routine groups (Χ ^2^= 6.0, *p*=0.196). Following adjustment for age, the proportion of HR-HPV positive attendees with an abnormal cytology result (N=618), with HPV16/18 positivity was 53.9% (95%CI: 34.8-73.1%) among the ineligible group, 18.1% (95%CI: 13.5-22.7%) among the catch-up-eligible group, and 6.5% (95%CI: 3.6-9.3%) among the routine-eligible group (Figure 2).

**Figure 2.**
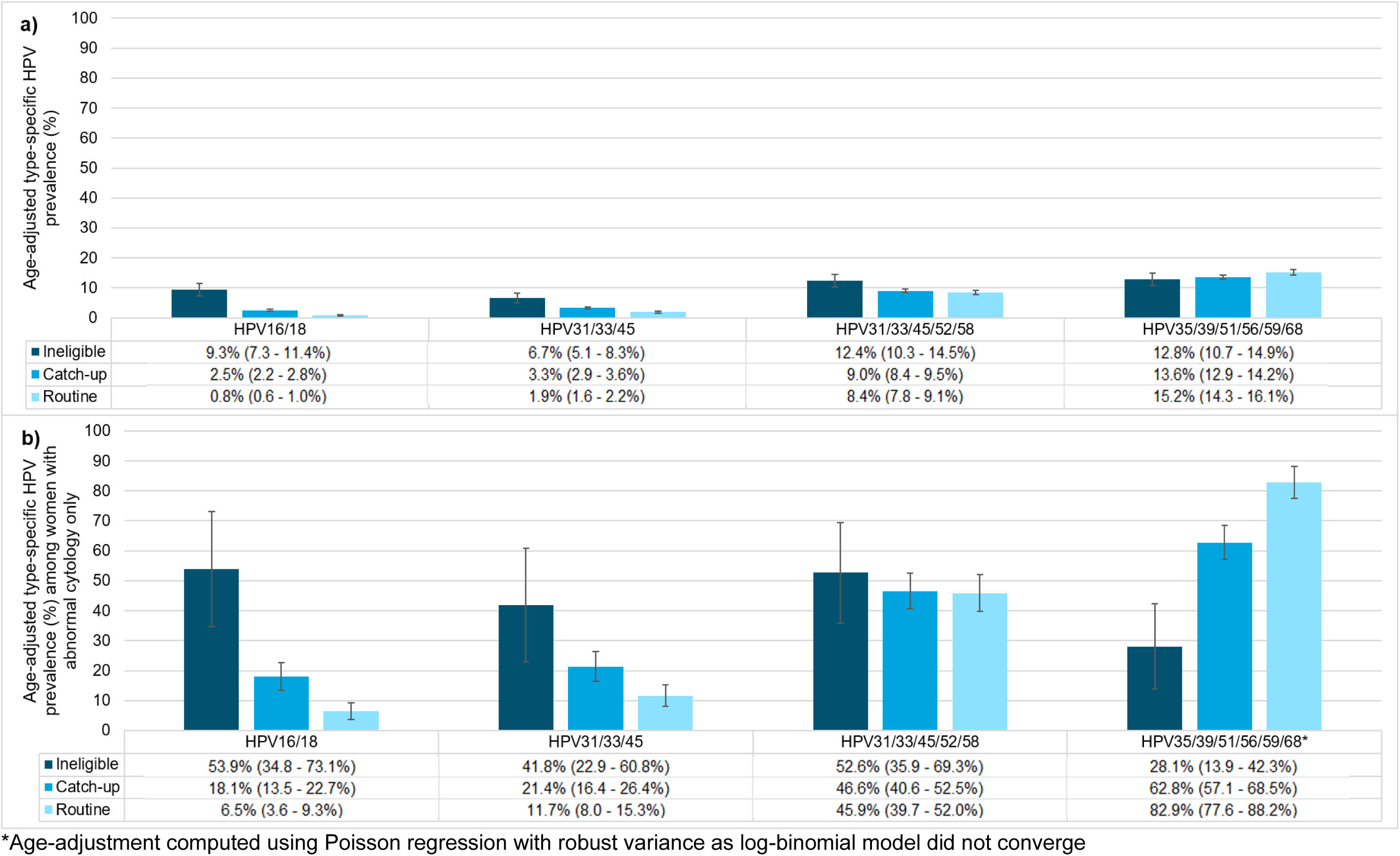
(a) Age-adjusted type-specific HPV prevalence estimates among all women sampled aged 25-29 from 2016-2025 (N=19,407) and (b) among only those with abnormal cervical cytology results following positive high-risk-HPV screening (N=618)

### Non-16/18 nonavalent vaccine HR types (HPV31/33/45/52/58)

Estimated age-adjusted prevalence of non-bivalent-vaccine-types HPV31 and HPV45 was 2.5% (95%CI: 1.4-3.6%) and 3.7% (95%CI: 2.4-4.9%) respectively among the ineligible group, 1.0% (95%CI: 0.8-1.2%) and 1.2% (95%CI: 1.0-1.4%) among the catch-up-eligible group, and 0.3% (95%CI: 0.2-0.5%) and 0.6% (95%CI: 0.4-0.7%) among the routine-eligible group, respectively (Table 3). Following adjustment for confounders, the adjusted prevalence of HPV31 was 85% (95%CI: 72-92%) lower among the routine-eligible group (aPR: 0.15, 95%CI: 0.08-0.28) and 55% (95%CI: 28-72%) lower among the catch-up-eligible group (aPR:0.45, 95%CI: 0.28-0.72) compared with the ineligible group, while the estimated HPV45 prevalence was 84% (95%CI: 73-90%) lower among the routine-eligible group (aPR: 0.16, 95%CI: 0.10-0.27) and 67% (95%CI: 51-77%) lower among the catch-up group (aPR: 0.33, 95%CI: 0.23-49; Figure 1; Table 4).

Estimated adjusted vaccine effectiveness was 99% (aPR 0.01; 95%CI: <0.01-0.05) against HPV31 and 93% (aPR: 0.07; 95%CI: 0.02-0.21) against HPV45 (Table 4). There was no evidence for any differences in prevalence estimates of HPV33 between the ineligible and routine groups (Table 4). Age-adjusted prevalence estimates of HPV52 and HPV58 ranged from 4.5-5.0% and 1.9-2.5% respectively (Table 3) and showed no evidence for any differences across vaccination eligibility groups, however had a positive association with first-dose vaccination coverage by cohort (Table 4). Among women with an abnormal cytology result, the age-adjusted proportion of samples positive for HPV31/33/45 types was 41.8% (95%CI: 22.9-60.8%) among the ineligible group, 21.4% (95%CI: 16.4-26.4%) among the catch-up group, and 11.7% (95%CI: 8.0-15.3%) among the routine-eligible group (Figure 2). The age-adjusted proportion of samples with HPV31/33/45/52/58 types present among women with abnormal cytology was 52.6% (95%CI: 35.9-69.3%) among vaccination-ineligible women, 46.6% (95%CI: 40.6-52.5%) among catch-up eligible women and 45.9% (95%CI: 39.7-52.0%) among routine-eligible women (Figure 2).

### Non-vaccine HR types (HPV35/39/51/56/59/68) and sexual risk indicator (HPV26/53/66/70/73)

There was no evidence of a change in any estimated prevalence of individual non-vaccine HR-HPV types 35, 39, 51, 56, 59 or 68 between vaccination-eligible and ineligible groups, however there was a negative association with HPV35 and first-dose vaccination coverage by cohort (aPR: 0.24 [95%CI: 0.08 - 0.76]; Table 4). The proportion of women with an abnormal cytology result who had a HPV35/39/51/56/59/68-positive sample increased from 28.1% (95%CI: 13.9-42.3%) in the ineligible group to 82.9% (95%CI: 77.6-88.2%) among the routine group (Figure 2). While presence of the sexual risk indicator (HPV26/53/66/70/73) decreased with age overall (3.2% [95%CI: 2.73-3.61%] among 25-year-olds; 1.9% [95%CI: 1.40-2.50%] among 29-year-olds [Χ ^2^= 30.9, *p*<0.001]), following adjustment for age distribution, presence of the sexual risk indicator (HPV26/53/66/70/73) did not differ between groups (ineligible: 3.0%, 95%CI: 0.2-4.2%, catch-up-eligible: 2.6%, 95%CI: 2.2-2.9%, routine-eligible: 2.6%, 95%CI: 2.2-2.9%).

### Sensitivity Analyses

If we assumed the same type-distribution in the excluded screen-positive samples (which had no HR type detected by our assay) as we found in the included screen-positives, this increased the overall HR-positivity rate from 19.0% to 22.7%. While the difference in prevalence estimates for individual HR-HPV types was within one percentage-point, these small differences compounded when presenting combined HR-HPV type groupings. The prevalence of HPV16/18 would have been around 8.9% among the vaccination-ineligible group (0.4% lower than in the primary analysis), around 4.1% among the catch-up group (1.5% higher than in the primary analysis) and 1.5% among the routine group (0.7% higher than in the primary analysis; Supplementary Table 1).

HR-HPV screening positivity ranged from 11.5%-28.7% between submitting laboratories. Not all laboratories submitted data each year of the collection, or for all vaccination cohorts (Table 2). In the second sensitivity analysis dataset (N=17,469), estimated age-adjusted prevalence ratios were slightly lower among the routine-eligible group versus the ineligible group, compared with the main dataset (Supplementary Table 2). HPV16/18 prevalence was 9.3% (95%CI: 7.3-11.4%) among the vaccination-ineligible group and 0.6% (95%CI: 0.4-0.8%) among the routine-eligible group. Age-adjusted HPV31 and HPV45 prevalence estimates were 2.6% (95%CI: 1.4-3.7%) and 3.8% (95%CI: 2.4-5.1%) respectively among the vaccination-ineligible group and 0.2% (95%CI: 0.1-0.4%) and 0.5% (95%CI: 0.3-0.6%) among the routine-eligible group respectively (Supplementary Table 2).

## Discussion

These data provide the first findings of bivalent vaccination impact among cervical screening attendees eligible for routine vaccination at age 12-13 in England. The prevalence of the vaccine-targeted HR-HPV types HPV16 and HPV18 among women aged 25-29 was 90% lower (95%CI: 86–93%) in those eligible for routine vaccination than in those ineligible for vaccination, falling to 0.8% (95%CI: 0.6–1.1%) in women eligible for vaccination at age 12–13. Given a first-dose bivalent HPV vaccination coverage of 55.6-81.9% among catch-up eligible and 85.9-90.6% among routine-eligible groups (13, 14), this suggests an estimated vaccine effectiveness of 97% (95%CI: 93-99%). These data present a dramatic shift in HPV16/18 epidemiology among cohorts eligible for bivalent vaccination, in line with other studies (19, 20). As the first line of defence against HPV-related disease, this impact is crucial for England in its progress towards the global strategy to eliminate cervical cancer (21). These declines in infection are so far consistent with models predicting a dramatic decline in cervical cancers following the vaccination programme introduction (22), and the substantial declines in cervical cancer rates that have already begun to materialise among young vaccination-eligible women (23, 24).

A baseline serology survey in England showed that, without vaccination, HR-HPV prevalence rose in the late teens, peaked in the early twenties, and then stabilised or declined towards age 29 (25). This echoes the risk profile of many common sexually transmitted infections, which peak at ages 18-24 in early years of sexual activity (26). In these data, prevalence of our indicator of sexual risk remained highest at age 25 and reduced with age. In contrast, while the crude prevalence of HPV16/18 declined with age in the vaccination-ineligible group, there was no longer a detectable association with age among the catch-up and routine eligible groups.

Age-adjusted HPV16/18 among vaccination-ineligible attendees was 9.3% (95%CI: 7.3 – 11.4%), dropping to 2.5% (95%CI: 2.2 - 2.8%) among the catch-up eligible group and 0.8% (95%CI: 0.6 - 1.0%) among the routine-eligible group. A similar pre-vaccination collection among women attending cervical screening from 2007-2009 estimated HPV16/18 prevalence at 9.2% (95%CI: 6.1–13.7%) (12), while baseline data collected from 24-25-year-old women attending the HR-HPV primary screening pilot in 2013 estimated HPV16/18 prevalence among vaccination-ineligible women at 11.8% (95%CI: 10.4%-13.4%) and 3.1% (95%CI: 2.8-3.4%) among catch-up-eligible women by 2018 (20). In Scotland, among vaccination-ineligible 20-21-year-old women attending cervical screening in 2009, HPV16/18 prevalence was 28.9% (95%CI: 26.7-31.1%), whereas HPV16/18 prevalence among same-aged vaccination-eligible women in 2015 was just 4.8% (95%CI: 3.8-5.9%) (27). Similarly large declines have also been observed in other countries with school-based vaccination achieving high coverage, such as Australia (28) and Sweden (29). Our findings validate the early HPV16/18 prevalence declines among catch-up cohorts and now present the first evidence of even larger declines in routine-eligible cohorts in England.

We observed strong evidence of cross-protective effects from the bivalent vaccine against HPV31 and HPV45, with a decline of 85% (95%CI: 72-92%) in HPV31 prevalence and a decline of 84% (95%CI: 73-90%) in HPV45 prevalence among the routine-eligible compared with the ineligible cohort groups respectively, in line with other literature (15, 18). There was no indication of cross-protective effects against HPV33, for which cross-protection has been evidenced in vaccine trials, however evidence has been less consistent in real-world vaccination impact studies (16, 30). In Scotland, by comparison, HPV31/33/45 prevalence declined from 14.2% (95%CI: 12.0-16.7%) in the baseline cohorts to 2.6% (95%CI: 1.9-3.6%) in vaccination-eligible cohorts (27).

Regarding other non-vaccine HR-HPV types, we saw some indication of an association between vaccination coverage by cohort and prevalence of both HPV52 and HPV58, despite there being no evidence of an increase in their prevalence between cohort groups. As there was such a strong negative association with first-dose vaccination coverage and HPV16/18 prevalence, this association in some non-vaccine types may be explained by viral unmasking whereby these genotypes may have been less detectable by the assay in HPV16/18 coinfections due to competition for molecular resources (31). There is some evidence of viral and clinical unmasking leading to increased detection of non-vaccine HR-HPV types, particularly HPV52(32), however, there remains little likelihood or evidence of type replacement (33). It will, though, be important to monitor non-vaccine type prevalence among cohorts eligible for the nonavalent vaccine to remain vigilant for these signals and develop understanding of HPV epidemiology in response to vaccine changes.

Among vaccination-eligible women with cytological abnormalities, our results show a proportional reduction in HPV16/18/31/45 when compared with vaccination-ineligible groups and with pre-vaccination baseline data (12). HPV16/18 was present in just 6.5% (95%CI: 3.6-9.3%) of samples from routine-eligible women, whereas other HR nonavalent vaccine types HPV31/33/45/52/58 were present in 45.9% (95%CI: 39.7-52.0%) of routine-eligible women, and non-nonavalent HR-HPV types HPV35/39/51/56/59/68 were present in 82.9% (95%CI: 77.6-88.2%) of routine-eligible women.

As the prevalence of the highest risk HPV types declines, the positive predictive value of the current HPV screening assay against CIN2+ is likely to reduce as less carcinogenic types make up a higher proportion of those tested (31). This will become more relevant as nonavalent vaccine-eligible women age into screening, having received additional direct protection against HR types HPV 31, 33, 45, 52, and 58 with a combined global population attributable factor of 17.7% of invasive cervical cancers, 10.1% of which is attributed to HPV33/52/58, types that are not associated with cross-protection from the bivalent vaccine (34). This may have implications for risk: benefit considerations around cervical screening among populations with high vaccination coverage in the future While NHSCSP screen-positives flagged due to slow-progressing HPV types should in most cases be mitigated by cytology triage, this could lead to unnecessary colposcopy and overtreatment of these cases.

Since July 2025, women and people with a cervix aged 25-49 attending screening in England who test negative for HR-HPV are invited for 5-year recall, a change to the previous 3-year interval enabled by switching to primary screening by HR-HPV testing (35). In light of declines in HR-HPV prevalence amongst vaccination-eligible women, some countries with high HPV vaccination uptake such as Australia (36) and the Netherlands (37) have extended screening intervals further for these women and have begun to offer self-collected samples (38). Our data may contribute to consideration of such changes in England.

This study has a number of limitations. Except for results with possible HR type HPV66 only, just 79% of screen-positive samples tested had a HR-HPV type-specific result. Discordance between our HR-HPV type-specific testing and screening results may be due to instability of the sample due to transport and storage prior to re-testing and reduced sensitivity of the type-specific in-house assay compared with the commercial assays used by NHSCSP laboratories. In total 1,154 screen-positive samples (including 994 samples without a type-specific result and 160 with only HPV66) were not included in the primary dataset analysed by HR-HPV type. While those excluded were similar to those included by age at attendance and submitting laboratory, more of these were collected in later years and therefore included more of the younger cohorts. While these samples were excluded from both the numerator and denominator in the primary analysis, the true type distribution among this sample remains unknown. An initial sensitivity analysis imputed missing data using adjusted prevalence estimates among the included HR-HPV positives. The individual prevalence of each HR-HPV type did not differ more than one percentage point between estimates. This difference is notable, however, when prevalence was very low, particularly among the routine group. The prevalence estimates presented in the primary analysis should therefore be interpreted with this in mind. Additionally, we assumed robust sensitivity of the NHSCSP HR-HPV screening to determine HR-HPV negative status, and we did not collect or test these samples.

There were also limitations to the data available and in sample collection. The data collected were for a sample of women tested through the national cervical screening programme by a handful of laboratories across England. More detailed demographic data were not available, so we could not assess representativeness by additional participant characteristics, nor could we assess relative representativeness of the datasets used for sensitivity analyses. While individual vaccination status was not available for direct calculation of vaccine effectiveness, these data do demonstrate real-world impact inclusive of likely considerable herd protection effects.

Laboratories were asked to provide a sample of their HR-HPV screening positive and negative data up to pre-defined targets intended to be representative of local throughput, however practical issues with sample and data collection may have compromised this. A sensitivity analysis was run to exclude data where these issues were identified, and findings were largely similar to those in the primary analysis. It was not possible to restrict attendances to routine screening or exclude recall attendances, meaning attendees with a HR-HPV-positive screening result may be slightly overrepresented, particularly during 2020 when cervical screening attendance was negatively impacted by the COVID-19 pandemic (39).

While women attending cervical screening are largely representative of the general population, they are by nature those engaged in care and are unlikely to include some of the women at greatest risk for poor HPV-related health outcomes. Data from NHSCSP attendees may provide an underestimate of the prevalence of HPV16/18 in the whole population. The high degree of protection against infection with vaccine-type HPV and HPV31/45, beyond expectations based on efficacy estimates, is also likely in part due to higher vaccination coverage among the cervical screening population than among the general population, as have been shown in Scotland (40). These results, though, provide additional context to the earlier evidence of large HPV16/18 prevalence declines among vaccinated and/or vaccination-eligible young women and girls aged 16-24 attending chlamydia screening in England (10, 41), which comprise a relatively higher risk population. Prevalence of HPV16/18 among 22-24-year-olds attending chlamydia screening was very high in the early years of the programme, at 16.4% (95%CI:14.3–18.5%) among ineligible cohorts in 2010–2011. This declined to 7.5% (95%CI: 2.7–12.3%) in 2014-2015 among ineligible and catch-up cohorts, to 1.7% (95%CI: 0.7-3.5%) in 2018-19 among catch-up and routine cohorts, and just 1% (95%CI: 0.0-5.3%) in 2020 (routine cohorts only) (11, 18), presenting a similar decline across comparable timeframes and cohort groupings to this study.

There is now extensive evidence demonstrating the impact of HPV vaccination of young girls in preventing HPV-related disease (24). The routine vaccination-eligible cohorts included in these data had high (>85%) vaccination coverage (13, 14), which was maintained up until 2020 when disruption associated with the COVID-19 pandemic negatively impacted vaccination uptake, and has not yet recovered (to end 2025) (42). The effects of this decline in direct and indirect protection at the population level may be moderated to some extent by the extension of the vaccination programme to include boys in 2019 (43), however strong inequalities in vaccination uptake by social deprivation level emerged alongside this decline (44). The effects of this on HPV prevalence and HPV-related disease are yet to be determined and will require close monitoring (45). Women living in deprived areas receive the most benefit from vaccination, (46), but are less likely to receive vaccination (44). In turn, unvaccinated women, who have higher rates of cytological abnormalities and cancers, are less likely to attend cervical screening (40). Uptake of both interventions is influenced by factors including level of socioeconomic deprivation and ethnicity, which may compound risk in the most deprived populations (47).

Significant declines in HR-HPV prevalence indicate a very strong protective effect of vaccination against HPV16 and HPV18 infection, and cross-protection against HPV31 and HPV45 among women eligible for the bivalent HPV vaccine. These results from over a decade of national surveillance data collection provide the first type-specific HPV prevalence estimates among routine vaccination-eligible women attending cervical screening in England. This demonstrates the success of the HPV vaccination programme against its primary objectives to reduce HR-HPV in this population. This ongoing surveillance has the power to monitor the real-world impact of policy changes such as gender-neutral vaccination and changes to vaccines in relevant vaccination cohorts as they enter the cervical screening programme. These data provide additional evidence of the impact of the national HPV vaccination programme in England and data to inform the NHSCSP amid a rapidly changing landscape. As nonavalent HPV vaccine types are expected to decline among the vaccination-eligible population, it will be important to continue monitoring non-vaccine HR-HPV types to understand their epidemiology and their role in HPV-related disease, particularly among those most at risk of poor health outcomes.

## Additional Information

## Data Availability

The datasets generated during and analysed during the current study are not publicly available due the protected nature of the data under UKHSA stewardship but are available by submitting an acceptable UKHSA data access request.

## Acknowledgements

We thank the UKHSA HPV laboratory staff (Victory Igbinosa, Kazutomo Yokoya and Dilan Patel), UKHSA staff involved in project administration (Manchari Rajkumar and Linda Gansberger) and staff involved in specimen and data collection at participating laboratories (including Gulseren Akgul and colleagues on behalf of Health Services Laboratories, London).

## Authors’ contributions

LS, MC, KS, KP and SB conceived this analysis. LS, MC and KS oversaw all surveillance work. AS, VF and BM contributed to sample and data collection. SB and KP conducted laboratory testing.

LS conducted data interpretation and statistical analysis, with support from LV, and wrote the first manuscript. All authors reviewed and contributed to successive drafts and approved the final version of the manuscript.

## Ethics approval and consent to participate

This surveillance was conducted as public health monitoring of the HPV vaccination programme: individual patient consent was not required. UKHSA has permission to handle these data for this purpose under Regulation 3 of The Health Service (Control of Patient Information) Regulations 2020 and Section 251 of the National Health Service Act 2006.

