## Supplementary Material for "Impact of HPV vaccination on high-risk HPV prevalence in 25-29-year-old women attending cervical screening in England 2016-2025"

**Supplementary Table 1:** Sensitivity Analysis #1. Estimated age-adjusted type-specific high-risk HPV prevalence among women attending for cervical screening, by vaccination eligibility 2016-2025 with prevalence among excluded HR-screen-positive data re-estimated using imputed number of positives.

|  | **Ineligible for vaccination** | | | **Catch-up eligible** | | | **Routine eligible** | | |
| --- | --- | --- | --- | --- | --- | --- | --- | --- | --- |
|  | *n* | *% (95%CI)* | *P_2_-P_1_^*^ (%)* | *n* | *% (95%CI)* | *P_2_-P_1_^*^ (%)* | *n* | *% (95%CI)* | *P_2_-P_1_^*^ (%)* |
| **Analysis dataset *(N=20,561)*** |  | *(N=1,245)* |  |  | *(N=11,699)* |  |  | *(N=7,617)* |  |
| Screen HR-HPV- / HPV66+ only | 913 | 73.3% (70.9% - 75.8%) | -1.5 | 9,137 | 78.1% (77.4% - 78.9%) | -2.9 | 5,834 | 76.6% (75.6% - 77.5%) | -5.5 |
| Typed/imputed HR-HPV+ | 332 | 26.7% (24.2% - 29.1%) | 0.1 | 2,562 | 21.9% (21.1% - 22.6%) | 2.9 | 1,783 | 23.4% (22.5% - 24.4%) | 5.5 |
| **Bivalent vaccine HPV types** |  |  |  |  |  |  |  |  |  |
| HPV16/18 | 111.1 | 8.9% (7.3% - 10.5%) | -0.4 | 477.7 | 4.1% (3.7% - 4.4%) | 1.5 | 115.1 | 1.5% (1.2% - 1.8%) | 0.7 |
| HPV16 | 87.0 | 7.0% (5.6% - 8.4%) | -0.7 | 389.5 | 3.3% (3.0% - 3.7%) | 1.3 | 92.7 | 1.2% (1.0% - 1.5%) | 0.6 |
| HPV18 | 24.3 | 1.9% (1.2% - 2.7%) | 0.2 | 106.7 | 0.9% (0.7% - 1.1%) | 0.4 | 21.6 | 0.3% (0.2% - 0.4%) | 0.1 |
| **Other nonavalent vaccine HR-HPV types** | | |  |  |  |  |  |  |  |
| HPV31/33/45/52/58 | 155.4 | 12.5% (10.6% - 14.3%) | 0.1 | 1,240.9 | 10.6% (10.0% - 11.2%) | 1.6 | 830.6 | 10.9% (10.2% - 11.6%) | 2.5 |
| HPV31/33/45 | 82.5 | 6.6% (5.2% - 8.0%) | -0.1 | 509.0 | 4.4% (4.0% - 4.7%) | 1.1 | 212.5 | 2.8% (2.4% - 3.2%) | 0.9 |
| HPV31 | 27.5 | 2.2% (1.4% - 3.0%) | -0.3 | 172.2 | 1.5% (1.3% - 1.7%) | 0.4 | 45.5 | 0.6% (0.4% - 0.8%) | 0.3 |
| HPV33 | 17.5 | 1.4% (0.8% - 2.1%) | 0.1 | 168.2 | 1.4% (1.2% - 1.7%) | 0.3 | 98.9 | 1.3% (1.0% - 1.6%) | 0.2 |
| HPV45 | 44.8 | 3.6% (2.6% - 4.6%) | -0.1 | 216.4 | 1.8% (1.6% - 2.1%) | 0.7 | 65.4 | 0.9% (0.7% - 1.1%) | 0.3 |
| HPV52 | 62.5 | 5.0% (3.8% - 6.2%) | 0.0 | 610.3 | 5.2% (4.8% - 5.6%) | 0.7 | 460.1 | 6.0% (5.5% - 6.6%) | 1.3 |
| HPV58 | 24.7 | 2.0% (1.2% - 2.8%) | 0.1 | 262.3 | 2.2% (2.0% - 2.5%) | 0.3 | 221.6 | 2.9% (2.5% - 3.3%) | 0.5 |
| **Other HR-HPV types** |  |  |  |  |  |  |  |  |  |
| HPV35/39/51/56/59/68 | 161.4 | 13.0% (11.1% - 14.8%) | 0.2 | 1,744.9 | 14.9% (14.3% - 15.6%) | 1.3 | 1,447.1 | 19.0% (18.1% - 19.9%) | 3.8 |
| HPV35 | 18.3 | 1.5% (0.8% - 2.1%) | 0.3 | 151.2 | 1.3% (1.1% - 1.5%) | 0.3 | 76.5 | 1.0% (0.8% - 1.2%) | 0.1 |
| HPV39 | 38.3 | 3.1% (2.1% - 4.0%) | -0.3 | 380.1 | 3.2% (2.9% - 3.6%) | 0.4 | 273.4 | 3.6% (3.2% - 4.0%) | 0.9 |
| HPV51 | 30.0 | 2.4% (1.6% - 3.3%) | -0.3 | 276.3 | 2.4% (2.1% - 2.6%) | 0.4 | 217.5 | 2.9% (2.5% - 3.2%) | 0.7 |
| HPV56 | 28.5 | 2.3% (1.5% - 3.1%) | -0.1 | 355.3 | 3.0% (2.7% - 3.3%) | 0.3 | 265.0 | 3.5% (3.1% - 3.9%) | 0.8 |
| HPV59 | 31.1 | 2.5% (1.6% - 3.4%) | -0.3 | 286.2 | 2.4% (2.2% - 2.7%) | 0.4 | 185.0 | 2.4% (2.1% - 2.8%) | 0.6 |
| HPV68 | 17.6 | 1.4% (0.8% - 2.1%) | 0.1 | 207.8 | 1.8% (1.5% - 2.0%) | 0.2 | 144.5 | 1.9% (1.6% - 2.2%) | 0.3 |
| **Multiple infections** |  |  |  |  |  |  |  |  |  |
| >1 HR-HPV type | 93.6 | 7.5% (6.1% - 9.0%) | -1.0 | 656.9 | 5.6% (5.2% - 6.0%) | 1.1 | 379.3 | 5.0% (4.5% - 5.5%) | 1.5 |
| >1 HR-HPV type including HPV16/18 | 48.2 | 3.9% (2.8% - 4.9%) | -0.8 | 210.6 | 1.8% (1.6% - 2.0%) | 0.8 | 37.4 | 0.5% (0.3% - 0.6%) | 0.3 |

*Difference between P_2_ (prevalence estimate in sensitivity analysis #1) and P_1_ (prevalence estimate in primary analysis [Table 3])

**Supplementary Table 2:** Sensitivity Analysis #2. Estimated age-adjusted type-specific high-risk HPV prevalence among women attending for cervical screening, by vaccination eligibility 2016-2025 with data excluded where yearly screen HR-positive were proportionally over or under sampled by laboratory.

|  | **Ineligible for vaccination** | | | **Catch-up eligible** | | | **Routine eligible** | | |
| --- | --- | --- | --- | --- | --- | --- | --- | --- | --- |
|  | *n* | *% (95%CI)* | *P_3_ -P_1_^*^ (%)* | *n* | *% (95%CI)* | *P_3_ -P_1_^*^ (%)* | *n* | *% (95%CI)* | *P_3_ -P_1_^*^ (%)* |
| **NHSCSP screening result *(N=18,422)*** | *(N=1,245)* | | | *(N=10,777)* | | | *(N=6,400)* | | |
| Screen HR-HPV+ (all) | 338 |  |  | 2,445 |  |  | 1,299 |  |  |
| Screen HR-HPV+ (excluded) | 56 |  |  | 551 |  |  | 346 |  |  |
| **Analysis dataset *(N=17,469)*** |  | *(N=1,189)* |  |  | *(N=10,226)* |  |  | *(N=6,054)* |  |
| Screen HR-HPV- | 907 | 74.5 (72.1 - 76.9) | -0.3 | 8,332 | 81.0 (80.2 - 81.7) | -0.1 | 5,101 | 85.6 (84.6 - 86.5) | 3.5 |
| Typed HR-HPV+ | 282 | 27.6 (24.6 - 30.5) | 1.0 | 1,894 | 19.2 (18.4 - 20.0) | 0.2 | 953 | 14.5 (13.6 - 15.4) | -3.4 |
| **Bivalent vaccine HPV types** |  |  |  |  |  |  |  |  |  |
| HPV16/18 | 94 | 9.8 (7.6 - 12.0) | 0.5 | 250 | 2.6 (2.3 - 2.9) | 0.0 | 39 | 0.6 (0.4 - 0.8) | -0.3 |
| HPV16 | 73 | 8.2 (6.1 - 10.2) | 0.5 | 198 | 2.1 (1.8 - 2.4) | 0.0 | 34 | 0.5 (0.3 - 0.6) | -0.2 |
| HPV18 | 21 | 1.7 (0.9 - 2.6) | 0.0 | 55 | 0.5 (0.4 - 0.7) | 0.0 | 6 | 0.1 (0.0 - 0.2) | -0.1 |
| **Other nonavalent vaccine HR-HPV types** | | |  |  |  |  |  |  |  |
| HPV31/33/45/52/58 | 132 | 12.7 (10.6 - 14.9) | 0.4 | 894 | 9.1 (8.5 - 9.6) | 0.1 | 464 | 7.1 (6.4 - 7.7) | -1.3 |
| HPV31/33/45 | 70 | 6.9 (5.2 - 8.6) | 0.2 | 329 | 3.4 (3.0 - 3.7) | 0.1 | 99 | 1.5 (1.2 - 1.8) | -0.4 |
| HPV31 | 23 | 2.6 (1.4 - 3.7) | 0.1 | 103 | 1.1 (0.9 - 1.3) | 0.1 | 17 | 0.2 (0.1 - 0.4) | -0.1 |
| HPV33 | 15 | 1.4 (0.6 - 2.1) | 0.1 | 115 | 1.1 (0.9 - 1.4) | 0.0 | 53 | 0.8 (0.6 - 1.1) | -0.2 |
| HPV45 | 38 | 3.8 (2.4 - 5.1) | 0.1 | 118 | 1.2 (1.0 - 1.4) | 0.0 | 30 | 0.5 (0.3 - 0.6) | -0.1 |
| HPV52 | 53 | 5.2 (3.8 - 6.6) | 0.2 | 447 | 4.5 (4.1 - 5.0) | 0.0 | 270 | 4.1 (3.6 - 4.6) | -0.7 |
| HPV58 | 21 | 2.0 (1.1 - 2.9) | 0.0 | 190 | 1.9 (1.6 - 2.2) | 0.0 | 132 | 2.0 (1.7 - 2.4) | -0.4 |
| **Other HR-HPV types** |  |  |  |  |  |  |  |  |  |
| HPV35/39/51/56/59/68 | 137 | 13.2 (11.0 - 15.3) | 0.4 | 1,352 | 13.7 (13.0 - 14.4) | 0.1 | 818 | 12.5 (11.7 - 13.4) | -2.7 |
| HPV35 | 16 | 1.2 (0.6 - 1.9) | 0.0 | 107 | 1.0 (0.8 - 1.2) | 0.0 | 48 | 0.8 (0.6 - 1.1) | 0.0 |
| HPV39 | 32 | 3.5 (2.3 - 4.8) | 0.1 | 274 | 2.9 (2.5 - 3.2) | 0.0 | 152 | 2.2 (1.8 - 2.5) | -0.5 |
| HPV51 | 25 | 2.7 (1.6 - 3.8) | 0.0 | 191 | 2.0 (1.7 - 2.3) | 0.0 | 118 | 1.7 (1.4 - 2.0) | -0.5 |
| HPV56 | 24 | 2.5 (1.5 - 3.5) | 0.1 | 269 | 2.8 (2.4 - 3.1) | 0.0 | 142 | 2.1 (1.7 - 2.4) | -0.6 |
| HPV59 | 26 | 2.9 (1.7 - 4.0) | 0.1 | 203 | 2.1 (1.8 - 2.4) | 0.1 | 105 | 1.5 (1.2 - 1.8) | -0.3 |
| HPV68 | 15 | 1.3 (0.6 - 2.0) | 0.0 | 162 | 1.6 (1.4 - 1.9) | 0.1 | 83 | 1.3 (1.0 - 1.6) | -0.2 |
| **Multiple infections** |  |  |  |  |  |  |  |  |  |
| >1 HR-HPV type | 78 | 8.7 (6.7 - 10.7) | 0.1 | 440 | 4.6 (4.2 - 5.0) | 0.1 | 204 | 2.9 (2.5 - 3.3) | -0.6 |
| >1 HR-HPV type including HPV16/18 | 40 | 4.9 (3.2 - 6.7) | 0.2 | 92 | 1.0 (0.8 - 1.2) | 0.0 | 14 | 0.2 (0.1 - 0.3) | 0.0 |

*Difference between P_3_ (prevalence estimate in sensitivity analysis #2) and P_1_ (prevalence estimate in primary analysis [Table 3])
